# Risk of thrombotic complications in influenza versus COVID-19 hospitalized patients

**DOI:** 10.1101/2020.12.18.20248265

**Authors:** Dutch COVID & Thrombosis Coalition (DCTC), MV Huisman, Milou AM Stals, Marco JJH Grootenboers, Coen van Guldener, Fleur HJ Kaptein, Sander JE Braken, Qingui Chen, Gordon Chu, Erik M van Driel, Antonio Iglesias del Sol, Evert de Jonge, K Merijn Kant, Fleur Pals, Myrthe MA Toorop, Suzanne C Cannegieter, Frederikus A Klok

## Abstract

**Background:** Whereas accumulating studies on COVID-19 patients report high incidences of thrombotic complications, large studies on clinically relevant thrombosis in patients with other respiratory tract infections are lacking. How this high risk in COVID-19 patients compares to those observed in hospitalized patients with other viral pneumonias such as influenza is unknown.

**Objectives:** To assess the incidence of venous and arterial thrombotic complications in hospitalized influenza patients as opposed to that observed in hospitalized COVID-19 patients.

**Methods:** Retrospective cohort study; we used data from Statistics Netherlands (study period: 2018) on thrombotic complications in hospitalized influenza patients. In parallel, we assessed the cumulative incidence of thrombotic complications – adjusted for competing risk of death - in patients with COVID-19 in three Dutch hospitals (February 24th - April 26th 2020).

**Results:** Of the 13.217 hospitalized influenza patients, 437 (3.3%) were diagnosed with thrombotic complications, versus 66 (11%) of the 579 hospitalized COVID-19 patients. The 30-day cumulative incidence of any thrombotic complication in influenza was 11% (95%CI 9.4-12) versus 25% (95%CI 18-32) in COVID-19. For venous thrombotic complications (VTE) and arterial thrombotic complications alone, these numbers were respectively 3.6% (95%CI 2.7-4.6) and 7.5% (95%CI 6.3-8.8) in influenza versus 23% (95%CI 16-29) and 4.4% (95%CI 1.9-8.8) in COVID-19.

**Conclusions:** The incidence of thrombotic complications in hospitalized influenza patients was lower than in hospitalized COVID-19 patients. This difference was mainly driven by a high risk of VTE complications in the COVID-19 patients admitted to ICU. Remarkably, influenza patients were more often diagnosed with arterial thrombotic complications.

**Essentials:**

- It is unknown how COVID-19 compares to patients with other virus infections regarding thrombosis.
- Hospitalized patients with influenza and COVID-19 were evaluated and compared to each other.
- 30-day cumulative incidence of thrombosis was lower in influenza (11%) than in COVID-19 (25%).
- Difference was mainly driven by a high risk of VTE in COVID-19 patients admitted to the ICU.

## Introduction

The clinical course of acute infections may become complicated by venous and arterial thrombotic disease.[1, 2] Precipitating factors for thrombotic complications in this setting include inflammation, activation of the coagulation system, immobilization, and diffuse intravascular coagulation (DIC).[1, 2] In light of this, international guidelines recommend pharmacological thromboprophylaxis in patients with infectious diseases if they are hospitalized.[3]

Respiratory viruses, including influenza, were already known to lead to a procoagulant state.[4-6] Moreover, previous studies showed an transient increase in the incidence of thrombotic vascular complications after respiratory tract infections.[1, 7] Still, to date it is largely unknown how often influenza infection leads to thrombotic complications, since only small case series yielding conflicting results have been published.[4, 8-10] Contrarywise, the COVID-19 pandemic has led to a large number of cohort studies on thrombotic complications in hospitalized COVID-19 patients. These studies showed that COVID-19 infections were characterized by a sometimes excessive activation of blood coagulation [11-13] and high incidences of thrombotic complications were reported in patients with COVID-19, aggravated by admittance to the Intensive Care Unit (ICU).[14-19] How this high incidence of thrombotic complications in COVID-19 patients compares to those observed in hospitalized patients with non-COVID-19 virus infections such as influenza is largely unknown.[20] This is especially relevant to understand the etiology of COVID-19 associated thrombosis.[21, 22] We evaluated the incidence of thrombotic complications in hospitalized patients with influenza as opposed to that in hospitalized patients with COVID-19, in an effort to better understand the epidemiology of COVID-19 associated thrombotic complications.

## Methods

### Setting and study population

We used anonymous information on registered diagnoses during hospital admissions in Dutch hospitals from the National Basic Register of Hospital Care of Dutch Hospital Data (DHD), which includes all general and academic Dutch hospitals, provided via Statistics Netherlands (“Centraal Bureau voor de Statistiek”, CBS), for information on hospitalized influenza patients. Patients hospitalized with an admission date between January 1st 2018 and November 30th 2018 with a diagnosis of influenza were included for the primary analysis and data from January 1^st^ 2013 until December 31^st^ 2018 were used to collect information on comorbidities. Information on the use of pharmacological thromboprophylaxis was not available in DHD.

We also used data from adult COVID-19 patients admitted to the wards and ICUs of one university hospital (Leiden University Medical Center, Leiden, the Netherlands) and two non-university teaching hospitals (Amphia Hospital Breda and Alrijne Hospital Leiderdorp, both in the Netherlands) between February 24^th^ and April 26^th^ 2020. COVID-19 was confirmed by a positive polymerase chain reaction (PCR) test or considered positive in patients with a negative PCR but highly suggestive symptoms and typical COVID-19 abnormalities on CT-scan of the chest (CO-RADS 4 or 5 following Dutch Radiology Society[23]) with no alternative diagnosis. All COVID-19 patients had received pharmacological thromboprophylaxis from admission on, according to local hospital protocols, which changed over time (Appendix 3). Informed consent was obtained by an opt-out approach. This study was approved by the Institutional Review Boards of the LUMC for observational studies.

### Objectives

The primary objective was to assess the incidence of venous and arterial thrombotic complications in hospitalized influenza patients (general ward and ICU combined) and compare this with that observed in our cohort of hospitalized COVID-19 patients.

### Data collection

A diagnosis of influenza in the allocated time frame was identified in the DHD data based on International Classification of Diseases (ICD) codes (Appendix 1). For those patients who had more than one influenza related hospitalization, only the first hospitalization was included. In these data, a differentiation is made between admission to a normal ward only or (temporarily) admission to an ICU, but it was not possible to distinguish exact days of admittance on the general ward versus ICU. Therefore, influenza patients in the DHD database were categorized as hospitalized at the ward or ICU by the following definition: 1. Ward: ‘hospitalizations that did not involve ICU admission’ (includes patients that were only admitted to the wards) and 2. ICU: ‘hospitalizations that did involve ICU admission’ (includes patients that were admitted to the ICU at some timepoint during hospitalization). Patient characteristics of included influenza patients as age, sex, and comorbidities were collected. A history of comorbidities as malignancy and prior history of venous thromboembolism (VTE) were screened for within 8 years before the influenza related hospitalization. Prior to 2018, categorization of the hospitalization as ward and/or ICU was not possible as this information was not available in the CBS database. Therefore, we studied the incidence of thrombotic complications in influenza related hospitalizations during influenza seasons between 2013 and 2017 as a sensitivity analysis, rather than as a main analysis. Influenza seasons started from the 40^th^ week of one year to the 20th week of the next year.

The patient charts from COVID-19 patients were retrospectively scrutinized for baseline characteristics and outcomes of interest. For COVID-19 we were able to strictly distinguish general ward from ICU admission by exact days of admittance, but we used the influenza categorization of ward and ICU in order to be consistent.

### Outcomes

Information on thrombotic complications in influenza was identified by ICD codes (Appendix 1). Any VTE (DVT and/or PE) or arterial thrombotic complication (ATE: ischemic stroke, myocardial infarction and/or systemic arterial embolism) that occurred during the index hospitalization was regarded an outcome event. The exact date of the thrombotic complication during admission was unavailable in DHD. Therefore, the date halfway the hospitalization period was chosen as the date of the thrombotic complication in influenza patients.

In COVID-19 patients, information on thrombotic complications was identified based on scrutinizing patient charts. Thrombotic complications consisted of acute pulmonary embolism (PE), deep-vein thrombosis (DVT), ischemic stroke, myocardial infarction and systemic arterial embolism. No VTE screening strategies were applied during this study in COVID-19 patients. Upon clinical suspicion of one of the thrombotic complications, appropriate diagnostic tests were applied, i.e. computed tomography pulmonary angiography (CTPA) for suspected PE and compression ultrasonography (CUS) for suspected DVT, cardiac enzymes including troponin, electrocardiogram and echocardiography for suspected acute coronary syndrome, and CT scan of the brain and CT angiography of the carotid and intracerebral arteries for suspected ischemic stroke.

### Statistical analysis

Patient characteristics were described using standard descriptive statistics. The analysis on the comparison between influenza and COVID-19 patients, comprised the full time of hospitalization (ward and ICU combined). Hospitalized influenza patients were followed from the day of hospital admission (i.e., the starting date) until day of discharge, day of death or end of data collection (31th December 2018), whichever came first. COVID-19 patients were followed from the day of hospital admission until discharge, transfer to another hospital, until they died, or until end of data collection (between April 17 and 26, 2020), whichever came first.

The incidence of thrombotic complications was estimated for all ICU and non-ICU patients combined, patients admitted to general wards only and for patients who had been admitted to the ICU at some timepoint during hospitalization (according to the DHD categorization of ward and ICU), and for all thrombotic complications as well as for venous and arterial complications separately. Cumulative incidences were estimated using the Kaplan-Meier method and the cumulative incidence competing risk (CICR) method, to adjust for the competing risk of death. In addition, incidence rates were calculated over the total follow-up period.

SPSS Statistics version 25.0 and R version 4.0.2 served for data analysis.

## Results

### Patients

A total of 13.217 hospitalized influenza patients and 579 hospitalized COVID-19 patients were included. Characteristics of both patient cohorts are summarized in Table 1. Patients with influenza and COVID-19 were comparable with regard to age, history of cancer, and length of hospital stay (median and IQR: five days (3-10) in influenza and seven days (4-11) in COVID-19). There were more male patients with COVID-19 than with influenza.

**Table 1:** Characteristics of hospitalized influenza patients and hospitalized COVID-19 patients

|  | Influenza<br>(N=13217)* | COVID-19<br>(N=579)* |
| --- | --- | --- |
| Age (mean, SD) | 69 (19) | 67 (13) |
| Male sex (number, %) | 6306 (48) | 380 (66) |
| Body weight in kg (median, IQR) | NA | 84 (73-95) |
| Cancer history (number, %)<sup>~</sup> | 2343 (18) | 86 (15) |
| Prior history VTE (number, %)<sup>~</sup> | 404 (3.1) | NA |
| Therapeutic anticoagulation at admission (number, %) | NA | 77 (13) |
| Length of hospital stay in days (median, IQR) | 5 (3-10) | 7 (4-11) |

Note: SD: standard deviation; IQR: interquartile range; VTE: venous thromboembolism; n: number; NA : not available
\* Ward and Intensive Care Unit patients combined
<sup>~</sup> Within influenza this could only be determined within 8 years before date of admission

### Thrombotic complications in influenza patients

Of the influenza patients, 437/13.217 patients (3.3%) were diagnosed with thrombotic complications, of whom 126 with venous and 319 with arterial complications. The 30-day cumulative incidence of all thrombotic complications, adjusted for competing risk of death, was 11% (95%CI 9.4-12) (Table 2).

**Table 2:** Cumulative incidences of thrombotic complications in hospitalized influenza patients versus hospitalized COVID-19 patients

|  | Influenza~<br>Crude<br>% (95% CI) | Influenza~<br>Adjusted^<br>% (95% CI) | COVID-19*<br>Crude<br>% (95% CI) | COVID-19*<br>Adjusted^<br>% (95% CI) |
| --- | --- | --- | --- | --- |
| All thrombotic complications |  |  |  |  |
| 10 days | 4.8% (4.3-5.4) | 4.6% (4.1-5.2) | 13% (9.3-17) | 12% (8.9-15) |
| 20 days | 8.9% (7.8-10) | 8.2% (7.3-9.2) | 30% (21-38) | 24% (18-30) |
| 30 days | 12% (10-14) | 11% (9.4-12) | 32% (22-41) | 25% (18-32) |
| VTE |  |  |  |  |
| 10 days | 1.3% (0.97-1.6) | 1.2% (0.95-1.5) | 12% (8.2-15) | 11% (7.7-14) |
| 20 days | 2.8% (2.2-3.5) | 2.6% (2.1-3.2) | 27% (19-36) | 21% (16-28) |
| 30 days | 4.1% (2.9-5.2) | 3.6% (2.7-4.6) | 29% (20-38) | 23% (16-29) |
| ATE |  |  |  |  |
| 10 days | 3.6% (3.2-4.1) | 3.5% (3.1-4.0) | 2.4% (0.83-4.0) | 2.2% (1.1-4.1) |
| 20 days | 6.3% (5.4-7.2) | 5.8% (5.0-6.7) | 3.9% (0.57-7.2) | 3.2% (1.4-6.4) |
| 30 days | 8.4% (6.9-10) | 7.5% (6.3-8.8) | 5.7% (0.8-11) | 4.4% (1.9-8.8) |

Note: All thrombotic complications: VTE and ATE together; VTE: venous thrombotic complications; ATE: arterial thrombotic complications; CI: confidence interval
~13217 influenza patients (ward and Intensive Care Unit combined)
\*579 COVID-19 patients (ward and Intensive Care Unit combined)
^Adjusted: cumulative incidence adjusted for competing risk of death

The 30-day adjusted cumulative incidence for venous thrombotic complications was 3.6% (95%CI 2.7-4.6) and for arterial thrombotic complications 7.5% (95%CI 6.3-8.8). The incidence-rates in hospitalized influenza patients were 1.6/patient-year (95%CI 1.4-1.7) for any thrombotic complication, 0.44/patient-year (95%CI 0.37-0.53) for venous thrombotic complications and 1.1/patient-year (95%CI 1.0-1.3) for arterial thrombotic complications. The incidences calculated for the year 2018 were comparable with previous years (2013-2017; Appendix 2).

For influenza patients that had only been admitted to the ward, the adjusted 30-day cumulative incidences for all thrombotic complications, venous thrombotic complications and arterial thrombotic complications were: 8.9% (95%CI 7.5-10), 2.4% (95%CI 1.8-3.2) and 6.6% (95%CI 5.4-8.0), respectively. From these ward admitted patients, 766 out of 12412 died during hospitalization (6.2%). In influenza patients that had been admitted to the ICU, the adjusted 30-day cumulative incidences for all thrombotic complications, venous thrombotic complications and arterial thrombotic complications were: 18% (95%CI 14-23), 7.7% (95%CI 5.0-11) and 11% (95%CI 7.9-15), respectively (Table 3). In this group of patients, 171 out of 805 died during hospital admission (21%).

**Table 3:** Adjusted cumulative incidences of thrombotic complications in ward and ICU Influenza and COVID-19 patients*

|  | Influenza ward<br>N=12412<br>% (95% CI) | COVID-19 ward<br>N=401<br>% (95% CI) | Influenza ICU<br>N=805<br>% (95% CI) | COVID-19 ICU<br>N=178<br>% (95% CI) |
| --- | --- | --- | --- | --- |
| All thrombotic complications |  |  |  |  |
| 10 days | 4.3% (3.8-4.9) | 4.7% (2.6-7.9) | 6.8% (5.1-8.9) | 21% (15-28) |
| 20 days | 7.2% (6.2-8.2) | 6.1% (3.1-11) | 13% (10-17) | 35% (27-44) |
| 30 days | 8.9% (7.5-10) | 6.1% (3.1-11) | 18% (14-23) | 36% (28-45) |
| VTE |  |  |  |  |
| 10 days | 1.1% (0.80-1.4) | 3.1% (1.4-5.9) | 2.5% (1.5-3.9) | 21% (15-27) |
| 20 days | 2.2% (1.7-2.8) | 4.4% (1.8-8.8) | 4.9% (3.2-7.1) | 33% (25-42) |
| 30 days | 2.4% (1.8-3.2) | 4.4% (1.8-8.8) | 7.7% (5.0-11) | 34% (26-43) |
| ATE |  |  |  |  |
| 10 days | 3.4% (2.9-3.9) | 1.7% (0.61-3.9) | 4.4% (3.0-6.1) | 2.5% (0.83-5.9) |
| 20 days | 5.2% (4.4-6.1) | 1.7% (0.61-3.9) | 8.7% (6.4-11) | 3.8% (1.3-8.5) |
| 30 days | 6.6% (5.4-8.0) | 1.7% (0.61-3.9) | 11% (7.9-15) | 5.3% (2.0-11) |

\*Following the classification of ward (hospitalizations that did not involve ICU) and ICU (hospitalizations that did involve ICU) as is done in the CBS hospitalization data
Note: All thrombotic complications: VTE and ATE together; VTE: venous thrombotic complications; ATE: arterial thrombotic complications; N= number; CI: confidence interval
7 Adjusted: cumulative incidence adjusted for competing risk of death

### Thrombotic complications in COVID-19 patients

Of the 579 COVID-19 patients, 66 patients (11%) were diagnosed with 70 thrombotic complications. PE was the most frequent thrombotic complication of all (n=54, 77%). The 30-day cumulative adjusted incidence of all thrombotic complications was 25% (95%CI 18-32) (Table 2). The 30-day adjusted cumulative incidence for venous thrombotic complications was 23% (95%CI 16-29) and for arterial thrombotic complications 4.4% (95%CI 1.9-8.8). Incidence-rates in COVID-19 patients were 5.1/patient-year (95%CI 3.9-6.4) for any thrombotic complication, 4.5/patient-year (95%CI 3.4-5.8) for venous thrombotic complications and 0.78/patient-year (95%CI 0.39-1.4) for arterial thrombotic complications.

For the COVID-19 patients that had only been admitted to the ward, the adjusted 30-day cumulative incidences for all thrombotic complications, VTE alone and ATE alone were: 6.1% (95%CI 3.1-11), 4.4% (95%CI 1.8-8.8) and 1.7% (95%CI 0.61-3.9), respectively. From these ward admitted patients, 66 out of 401 died during hospitalization (17%). In COVID-19 patients that had been admitted to the ICU at any moment during the in-hospital stay, the adjusted 30-day cumulative incidences for all thrombotic complications, VTE alone and ATE alone were: 36% (95%CI 28-45), 34% (95%CI 26-43) and 5.3% (95%CI 2.0-11), respectively (Table 3). In this group of patients, 45 out of 178 died during hospital admission (25%).

## Discussion

In this study we observed that, compared to hospitalized influenza patients, patients admitted with COVID-19 had an distinctly increased risk for getting thrombotic complications by day 30. This risk was driven by a difference in venous thrombotic complications (3.6% in influenza and 23% in COVID-19) and was particularly observed in patients admitted to the ICU.

How often influenza infection led to clinically relevant thrombotic disease was hitherto underreported, since only small case series have been published, yielding conflicting results.[4, 8-10] Nevertheless, respiratory viruses, including influenza, were already known to lead to a procoagulant state.[4-6] Previous studies for instance showed a transient increase in the risk of vascular complications after respiratory tract infections, with a twofold increase for venous thrombotic disease and an even fivefold increase for ischemic heart disease.[1, 7] Of note, in our study arterial thrombotic complications, and in particular myocardial infarction, occurred more in influenza patients than in COVID-19 patients. A recent study showed stroke rates quite similar to our study.[24] Strokes occurred in 0.2% in the influenza patients and in 1.6% in the COVID-19 patients [24], in our study this was 0.8% in the influenza patients and 1.7% in the COVID-19 patients.

Furthermore, another Chinese study was recently published, which compared the incidence of VTE in patients hospitalized with COVID-19 versus those hospitalized with community-acquired pneumonia (CAP).[20] Surprisingly, this study found no difference in VTE rates between CAP and COVID-19.

However, COVID-19 patients in this study were overall younger and had fewer comorbidities.[20] In addition, arterial thrombotic complications were not included in this study and VTE risk is known to be lower in East Asians than in white Caucasians, which limits generalizing these findings to other populations. A recent large cross-sectional study on 80.261 influenza patients illustrated that the high risk of arterial thrombotic complications coincides with influenza virus infection.[25] Therefore, it is reasonable to believe that viral pneumonias such as influenza virus and SARS-CoV-2 virus both interact with the coagulation system.[4]

Virus specific features could play an important role and maybe explain the observed difference in thrombotic complications between influenza and COVID-19 patients.[4, 26, 27] Recent studies suggest the possibility of in-situ immunothrombosis in COVID-19 patients, due to a SARS-CoV-2 specific effect. Following this hypothesis, COVID-19 associated alveolar injury and an extreme inflammatory response contribute to small vessel thrombus formation in the lungs. Autopsy studies support this hypothesis by reporting the presence of platelet-fibrin thrombi in small arterial vessels of the lungs.[22, 26, 28] On the other hand, other studies observed a high incidence of DVT upon screening as well, which is compatible with the conventional thromboembolic origin of PE.[29, 30] Therefore, both mechanisms likely play a role in the pathophysiology of COVID-19 associated thrombosis and further research in this field is needed.

Methodological strengths of our study include the large sample size and multicenter study design. In addition, VTE screening strategies have not been applied during the course of this study and diagnostic tests were only performed upon clinical suspicion. Limitations of our study regarding influenza data are that not all baseline patient characteristics were available, since these could only be determined during a specific time period before hospital admission, and that exact dates of thrombotic complications were not available. Furthermore, we do not know whether, and if so, which dose thrombosis prophylaxis was given to the hospitalized influenza patients. National guidelines in the Netherlands recommend pharmacological thromboprophylaxis with low molecular weight heparin (LMWH) in ward admitted patients in the case of a PADUA Prediction Score ≥ 4 and in the absence of contra-indications. All patients admitted to the ICUs receive pharmacological thromboprophylaxis, irrespective of the PADUA Prediction Score.[31] Standard practice in the LUMC, Amphia Hospital and Alrijne Hospital is the administration of nadroparin 2850 IU sc. per day. It is likely that these recommendations were followed for the hospitalized influenza patients, but still we do not know for sure. In addition, outcome classification differed between influenza and COVID-19 patients, as information on thrombotic complications was identified in influenza based on ICD codes and in COVID-19 by patient chart review. Besides, it is important to recall that thrombotic risks in influenza may differ between the seasonal influenza virus – from which patients were included in this study - and the H1N1 influenza virus pandemic from the year 2009/2010. For COVID-19 data, limitations include the different doses of thrombosis prophylaxis being prescribed over time and the lowering threshold to suspect VTE over the course of time in COVID-19 patients. In addition, thrombosis in COVID-19 due to in-situ immunothrombosis could have been underestimated, since this diagnosis is often only established after death in pathology reports. Still we were interested in clinically relevant confirmed thrombosis and believe that this underestimation will likely be minimal, especially given this lowering threshold to suspect VTE in COVID-19. Furthermore, compared to influenza patients, we observed a higher mortality rate in the COVID-19 ward admitted patients, which indicates that the COVID-19 ward admitted patients had more severe illness than the influenza ward admitted patients. Although this could (partially) explain the observed difference in thrombotic risk between the ward admitted patients in both groups, this cannot explain differences found in thrombotic complications between patients that had been admitted to ICU, as mortality rates in these patients were quite comparable, but a large difference was found in (especially venous) thrombotic complications. Altogether, the comparison between influenza and COVID-19 patients could have been biased because of the difference in outcome classification and the lowered threshold for diagnostic testing upon suspicion of VTE in COVID-19 patients was probably lower than in influenza patients after the high incidence of thrombotic complications in COVID-19 was elucidated.

In conclusion, this study suggests that the incidence of thrombotic complications in hospitalized influenza patients is lower than in hospitalized COVID-19 patients. The difference in thrombotic complications in our study was particularly driven by an exceptionally high risk for VTE in the COVID-19 patients that had been admitted to ICU. Remarkably, influenza patients were more often diagnosed with arterial thrombotic complications than with VTE. Further studies are needed to substantiate our findings and to explore explanations for this difference.

## Supporting information

Appendix A: contributors DCTC

STROBE checklist

## Data Availability

Anonymous data on hospitalized COVID19 patients as collected in this study can be made available to others, on request to the corresponding author of this manuscript (MV Huisman). The request will be granted after approval of a methodologically sound proposal and a signed data sharing agreement is required. Data will be available after publication of the article and until five years following article publication. Unfortunately, data on hospitalized influenza patients cannot be shared with third parties as Statistics Netherlands does not permit this. No other relevant documents are available.

## The Dutch COVID & Thrombosis Coalition study group

### Authors

MAM Stals, MD: Department of Thrombosis and Hemostasis, Leiden University Medical Center, Leiden, the Netherlands. Gathered and verified data, performed the analyses and primarily drafted the first version of the manuscript.

MJJH Grootenboers, MD: Department of Pulmonology, Amphia Hospital Breda, the Netherlands. Gathered data and revised the manuscript critically for important intellectual content.

C van Guldener, MD: Department of Internal Medicine, Amphia Hospital Breda, the Netherlands. Revised the manuscript critically for important intellectual content.

FHJ Kaptein, MD: Department of Thrombosis and Hemostasis, Leiden University Medical Center, Leiden, the Netherlands. Gathered data and revised the manuscript critically for important intellectual content.

SJE Braken: Department of Thrombosis and Hemostasis, Leiden University Medical Center, Leiden, the Netherlands. Gathered data and revised the manuscript critically for important intellectual content.

Q Chen, MD: Department of Clinical Epidemiology, Leiden University Medical Center, Leiden, the Netherlands. Analysed the DHD Influenza data and revised the manuscript critically for important intellectual content.

G Chu, MD: Department of Thrombosis and Hemostasis, Leiden University Medical Center, Leiden, the Netherlands. Gathered data and revised the manuscript critically for important intellectual content.

EM van Driel, MD: Department of Intensive Care Medicine, Alrijne Hospital Leiderdorp, the Netherlands. Revised the manuscript critically for important intellectual content.

A Iglesias del Sol, MD: Department of Internal Medicine, Alrijne Hospital Leiderdorp, the Netherlands. Revised the manuscript critically for important intellectual content.

E de Jonge, MD: Department of Intensive Care Medicine, Leiden University Medical Center, Leiden, the Netherlands. Revised the manuscript critically for important intellectual content.

KM Kant, MD: Department of Intensive Care Medicine, Amphia Hospital Breda, the Netherlands. Revised the manuscript critically for important intellectual content.

F Pals: Department of Thrombosis and Hemostasis, Leiden University Medical Center, Leiden, the Netherlands. Gathered data and revised the manuscript critically for important intellectual content.

MMA Toorop, MD: Department of Clinical Epidemiology, Leiden University Medical Center, Leiden, the Netherlands. Analysed the DHD Influenza data and revised the manuscript critically for important intellectual content.

SC Cannegieter, PhD: Department of Thrombosis and Hemostasis, Leiden University Medical Center, Leiden, the Netherlands; Department of Clinical Epidemiology, Leiden University Medical Center, Leiden, the Netherlands. Designed the study and provided methodological input. In addition, revised the manuscript critically for important intellectual content.

FA Klok, MD: Department of Thrombosis and Hemostasis, Leiden University Medical Center, Leiden, the Netherlands. Designed the study, performed the analyses and drafted the first version of the manuscript and revised the manuscript critically for important intellectual content.

MV Huisman, MD: Department of Thrombosis and Hemostasis, Leiden University Medical Center, Leiden, the Netherlands. Designed the study, verified data, performed the analyses and drafted the first version of the manuscript and revised the manuscript critically for important intellectual content.

All authors agree with the final version.

## Contributors

See **Appendix A**.

## Acknowledgements

The authors thank Statistics Netherlands for providing data from the Dutch Hospital Data registry.

## Data sharing

Anonymous data on hospitalized COVID-19 patients as collected in this study can be made available to others, on request to the corresponding author of this manuscript (MV Huisman). The request will be granted after approval of a methodologically sound proposal and a signed data sharing agreement is required. Data will be available after publication of the article and until five years following article publication. Unfortunately, data on hospitalized influenza patients cannot be shared with third parties as Statistics Netherlands does not permit this. No other relevant documents are available.

