## Supplementary material for "Risk of thrombotic complications in influenza versus COVID-19 hospitalized patients"

**Appendix 1:** ICD codes of the studied variables in DHD database on influenza patients

| | ICD codes |
|---|---|
| Influenza | ICD-9: 487<br>ICD-10: J09, J10 |
| VTE | |
| DVT | ICD-9: 4511, 45111, 45119, 4512, 4518, 45181, 45189, 4519, 452, 4531, 4532, 4533, 4538, 4539<br>ICD-10: I801, I802, I803, I808, I809, I821, I822, I823, I828, I829 |
| PE | ICD-9: 4151<br>ICD-10: I26 |
| ATE | |
| Ischemic stroke | ICD-10: I63, I64, H341 |
| Myocardial infarction | ICD-10: I21, I22 |
| Systemic arterial embolism | ICD-10: I74 |
| TE | VTE + ATE |
| Malignant tumor | ICD-9: 14, 15, 16, 17, 18, 19, 20<br>ICD-10: C |
| DIC | ICD-9: 2866<br>ICD-10: D65 |

**Note:** ICD-9 codes were only used for dates before 2013.

Abbreviations: VTE, venous thromboembolism; DVT, deep vein thrombosis; PE, pulmonary embolism; ATE, arterial thromboembolism; TE, thromboembolism; DIC, disseminated intravascular coagulation.

**Appendix 2:** Cumulative incidences of thrombotic complications in the studied influenza patients (2013-2017)

| Outcome | 2013 | 2013 | 2014 | 2014 | 2015 | 2015 | 2016 | 2016 | 2017 | 2017 |
|---|---|---|---|---|---|---|---|---|---|---|
| | KM | CICR | KM | CICR | KM | CICR | KM | CICR | KM | CICR |
| | (%, 95% CI) | (%, 95% CI) | (%, 95% CI) | (%, 95% CI) | (%, 95% CI) | (%, 95% CI) | (%, 95% CI) | (%, 95% CI) | (%, 95% CI) | (%, 95% CI) |
| **Combined** | | | | | | | | | | |
| 10 days | 3.3% (2.2-4.5) | 3.4% (2.3-4.7) | NA | NA | 4.9% (3.9-6.0) | 5.0% (4.0-6.2) | 3.8% (2.9-4.6) | 3.8% (3.0-4.8) | 5.0% (4.2-5.8) | 5.1% (4.3-6.0) |
| 20 days | 6.3% (4.1-8.4) | 6.7% (4.5-9.4) | NA | NA | 7.6% (5.8-9.3) | 8.1% (6.3-10) | 7.4% (5.9-8.9) | 8.1% (6.4-10) | 8.0% (6.7-9.4) | 8.9% (7.4-10) |
| 30 days | 7.4% (4.7-10) | 8.4% (5.4-12) | NA | NA | 8.9% (6.7-11) | 10% (7.6-14) | 9.8% (7.8-12) | 12% (9.2-15) | 9.5% (7.9-11.2) | 12% (9.4-14) |
| **VTE** | | | | | | | | | | |
| 10 days | NA | NA | NA | NA | 1.8% (1.2-2.5) | 1.9% (1.3-2.6) | 1.4% (0.89-1.9) | 1.4% (0.98-2.0) | 1.5% (1.1-2.0) | 1.6% (1.2-2.1) |
| 20 days | 1.6% (0.45-2.8) | 1.8% (0.75-3.6) | NA | NA | 2.8% (1.7-3.9) | 3.1% (2.0-4.5) | 3.2% (2.1-4.3) | 3.6% (2.5-5.0) | 2.4% (1.7-3.1) | 2.5% (1.9-3.4) |
| 30 days | 2.3% (0.55-3.9) | 2.8% (1.0-6.1) | NA | NA | 3.5% (2.1-4.9) | 4.0% (2.5-6.2) | 4.8% (3.3-6.3) | 6.0% (4.1-8.3) | 3.1% (2.1-4.0) | 3.9% (2.5-5.7) |
| **ATE** | | | | | | | | | | |
| 10 days | 2.4% | 2.4% | NA | NA | 3.2% | 3.2% | 2.4% | 2.4% | 3.5% | 3.6% |

| | | | | | | | | | | |
|---|---|---|---|---|---|---|---|---|---|---|
| | (1.4-3.4) | (1.6-3.6) | | | (2.3-4.1) | (2.4-4.2) | (1.7-3.1) | (1.8-3.2) | (2.8-4.2) | (2.9-4.3) |
| 20 days | 4.6%<br>(2.8-6.5) | 4.9%<br>(3.2-7.2) | NA | NA | 4.8%<br>(3.4-6.2) | 5.1%<br>(3.7-6.8) | 4.9%<br>(3.6-6.2) | 5.3%<br>(4.0-6.9) | 5.8%<br>(4.7-7.0) | 6.5%<br>(5.2-8.0) |
| 30 days | 5.8%<br>(3.4-8.2) | 6.6%<br>(4.0-10) | NA | NA | 5.5%<br>(3.8-7.2) | 6.4%<br>(4.2-9.1) | 5.7%<br>(4.2-7.2) | 6.6%<br>(4.9-8.6) | 6.7%<br>(5.3-8.0) | 8.1%<br>(6.2-10) |

* considering competing risk

Abbreviations: KM: Kaplan-Meier estimator; CICR: cumulative incidence competing risk; CI: confidence interval; VTE, venous thromboembolism; ATE, arterial thromboembolism; NA: not available

Note: events<10 are not made available in CBS exports, 2014 results were not available due to a DHD/CBS storage error in that year

**Appendix 3:** Local hospital protocols for thrombosis prophylaxis in participating centers for COVID-19 patients.

| | **Ward** | **ICU** |
|---|---|---|
| Leiden University Medical Center | Nadroparin 2850 IU sc per day or 5700 IU per day if body weight >100 kg | Nadroparin 2850 IU per day or 5700 IU per day if body weight >100kg; nadroparin 5700 IU per day or 5700 IU twice daily if body weight >100 kg from April $10^{th}$ 2020 and onwards |
| Alrijne Hospital Leiderdorp | Nadroparin 2850 IU per day or 5700 IU per day if body weight >100 kg | Nadroparin 5700 IU per day or 5700 IU twice daily if body weight >100 kg |
| Amphia Hospital Breda | Nadroparin 2850 IU sc per day or 5700 IU per day if body weight >100 kg; nadroparin 5700 IU sc per day in all ward patients from March $30^{th}$ 2020 and onwards (twice daily if body weight >100 kg) | Nadroparin 2850 IU sc per day or 5700 IU per day if body weight > 100 kg; nadroparin 5700 IU sc per day in all ICU patients from March 30th 2020 onwards; nadroparin 5700 IU twice daily or nadroparin 7600 IU twice daily dependent on body weight from halfway April 2020 and onwards |

Note: IU: international units; sc: subcutaneous; kg: kilograms
