## Appendix A: contributors DCTC for "Risk of thrombotic complications in influenza versus COVID-19 hospitalized patients"

**Appendix A: CONSORTIUM MEMBERS Dutch COVID & Thrombosis Coalition**

Amsterdam University Medical Center:

- Location AMC:

Dr. M. Coppens

Prof. dr. N.P. Juffermans

Prof. dr. S. Middeldorp

- Location VUMC:

Prof. dr. C.M.P.M. Hertogh

J.G. Hugtenburg

Dr. E.J. Nossent

Erasmus Medical Center:

Drs. J. van den Akker

Dr. R. Bierings

Dr. H. Endeman

Dr. M. Goeijenbier

Prof. dr. D.A.M.P.J. Gommers

Prof. dr. M.P.G. Koopmans

Prof. dr. T. Kuiken

T. Langerak

Dr. M.N. Lauw

Prof. dr. M.P.M. de Maat

D. Noack

M.S. Paats

M.P. Raadsen

Dr. B. Rockx

Dr. C. Rokx

Dr. C.A.M. Schurink

K. Tong-Minh

Dr. L van den Toorn

Dr. C.A. den Uil
C. visser

Farmadam pharmacy:

Tineke Roest

Institute of Research of Hospital de la Santa Creu i Sant Pau, Barcelona:

Dr. J. Manuel Soria

Leiden University Medical Center:

M.L. Antoni

Dr. M. Bos

Drs. Burggraaf

Prof. S.C. Cannegieter

Prof. dr. H.C.J. Eikenboom

Dr. P.L. den Exter

Dr. J.J.M. Geelhoed

Prof. dr. M.V. Huisman

Prof. E. de Jonge

Drs. F.H.J. Kaptein

Dr. F.A. Klok

Dr. L.J.M. Kroft

Drs. L. Nab

Dr. M.K. Ninaber

Prof. dr. H. Putter

Dr. A.M. da Rocha Rondon

Dr. A.H.E. Roukens

Drs. M.A.M. Stals

Prof. dr. H.H. Versteeg

Dr. H.W. Vliegen

Dr. B.J.M. van Vlijmen

Maastricht University Medical Center:

Dr. B.C.T. van Bussel.

Prof. dr. T.M. Hackeng

Drs. T. van de Berg

Prof. dr. H. ten Cate

Dr. ir. Y. Henskens

Dr. H. Spronk

Prof. dr. L. Schurgers

Drs. R. Bruggemann

Dr. B. Spaetgens

Dr. K. Winckers

Drs. R. Olie

Drs. M. Mulder.

Drs. A. Hulshof.

Prof. dr. M.A. Spruit

Radboud University Medical Center:

Dr. J. Leentjens

Dr. Q. de Mast

Sanquin Research, Amsterdam:

Dr. M. van den Biggelaar

Prof. dr. J.C.M. Meijers

Prof. dr. J. Voorberg

Synapse Research Institute:

Dr. B. de Laat, biochemicus

Thrombosis services Maastricht:

Dr. A. Ten Cate-Hoek

University Medical Center Groningen:

Prof. dr. T. Lisman

Prof. dr. K. Meijer

University Medical Center Utrecht:

O.L. Cremer

Dr. G. Geersing

Prof dr. H.A.H. Kaasjager

N. Kusadasi

A. Huisman

Dr. M. Nijkeuter

Prof. dr. R.E.G. Schutgens

Dr. R.T. Urbanus

J. Westerink
